# Characterization of pediatric urinary microbiome at species-level resolution indicates variation due to sex, age, and urologic history

**DOI:** 10.1101/2024.05.16.24307309

**Authors:** Maryellen S Kelly, Erin M Dahl, Layla Jeries, Tatyana A Sysoeva, Lisa Karstens

## Abstract

**Background:** Recently, associations between recurrent urinary tract infections (UTI) and the urinary microbiome (urobiome) composition have been identified in adults. However, little is known about the urobiome in children. We aimed to characterize the urobiome of children with species-level resolution and to identify associations based on UTI history.

**Study design:** Fifty-four children (31 females and 21 males) from 3 months to 5 years of age participated in the study. Catheterized urine specimens were obtained from children undergoing a clinically indicated voiding cystourethrogram. To improve the analysis of the pediatric urobiome, we used a novel protocol using filters to collect biomass from the urine coupled with synthetic long-read 16S rRNA gene sequencing to obtain culture-independent species-level resolution data. We tested for differences in microbial composition between sex and history of UTIs using non-parametric tests on individual bacteria and alpha diversity measures.

**Results:** We detected bacteria in 61% of samples from 54 children (mean age 40.7 months, 57% females). Similar to adults, urobiomes were distinct across individuals and varied by sex. The urobiome of females showed higher diversity as measured by the inverse Simpson and Shannon indices but not the Pielou evenness index or number of observed species (p = 0.05, p=0.04, p = 0.35, and p = 0.11, respectively). Additionally, several species were significantly overrepresented in females compared to males, including those from the genera *Anaerococcus, Prevotella,* and *Schaalia* (p = 0.03, 0.04, and 0.02, respectively). Urobiome diversity increased with age, driven mainly by males. Comparison of children with a history of 1, 2, or 3+ UTIs revealed that urobiome diversity significantly decreases in the group that experienced 3+ UTIs as measured by the Simpson, Shannon, and Pielou indices (p = 0.03, p = 0.05, p = 0.01). Several bacteria were also found to be reduced in abundance.

**Discussion:** In this study, we confirm that urobiome can be identified from catheter-collected urine specimens in infants as young as 3 months, providing further evidence that the pediatric bladder is not sterile. In addition to confirming variations in the urobiome related to sex, we identify age-related changes in children under 5 years of age, which conflicts with some prior research. We additionally identify associations with a history of UTIs.

**Conclusions:** Our study provides additional evidence that the pediatric urobiome exists. The bacteria in the bladder of children appear to be affected by early urologic events and warrants future research.

## Introduction

The lower urinary tract of an adult human is inhabited by a microbiome composed of bacteria, fungi, and viruses that are associated with several urological conditions, including nephrolithiasis, urinary incontinence, urinary urgency, and bladder overactivity.[1–4] Evidence also suggests that the presence of specific microbes and their abundance may affect the development and progression of urinary tract infections (UTI).[1,2,5,6]) Several species of the adult urinary microbiome were identified as protective against uropathogens *in vitro* and in animal models.[5–7] It remains unknown how the urobiome is initially colonized, how it develops during childhood, and if early urobiome development and changes can lead to a predisposition to recurrent UTIs.

The development of this microbial niche after birth and through childhood has not been characterized in detail due to several obstacles. One complication is with the feasibility and ethical concerns in collecting urine samples aseptically from younger children with catheters or suprapubic aspiration. The current standard of urobiome urinary collection uses catheterized urine, but this method is deemed too risky to be approved for children who are not undergoing catheterization for a medically directed purpose.[8] The alternative to this is obtaining voided clean-catch urine, which is complicated by the transition of children from the diapered stage to the toilet-trained stage, leaving younger children hard to sample. A second complication is that children have smaller bladder capacities than adults.[9–11] Thus, it has been assumed that children’s urobiome microbial abundance will be smaller and more difficult to detect. Kassiri et al reported microbial load in pediatric males, which was on average less when compared to the adult male urobiome[13].

Despite these complications, there have been seven pediatric urobiome studies within the last four years that discuss the development of the urobiome from ∼2 weeks to 18 years of age for both sexes.[12–18] There is little overlap among the subgroups analyzed and the methods used (**Table S1**). Most studies used routine short amplicon 16S rRNA gene sequencing to record bacterial urobiome composition. Two of these reports also used enhanced quantitative urine culture (EQUC) or a custom modification of this to isolate species from pediatric urine samples, thus providing species-level resolution of culturable microbes.[16, 17] This improved taxonomic resolution is critical for understanding urobiome functioning, as identified species may be relevant in discovering any predisposition for UTI.

In this pilot study, we sought to improve the resolution of the compositional analysis of the pediatric urobiome. We implemented methods of microbiome collection from other well-studied low-biomass niches and collected urine-suspended cells via filtering.[19–21] In addition, we used a synthetic long-read sequencing method to establish better taxonomic resolution of bacteria based on full-length 16S rRNA gene sequences.[22] Using this protocol, we analyzed bacterial composition of the urobiome to the species level in infants and young children. We detected differences in microbe diversity and presence by age, sex, and history of UTI.

## Methods

### Clinical population

The research study was approved by Duke University Medical Center Institutional Review Board under protocol #Pro0010031. From 2018-2021 children from birth to age 5 years who had a voiding cystourethrogram (VCUG) conducted for a clinical indication were offered enrollment. At baseline, parents or guardians (hereafter “parents”) granted permission for a urine sample to be collected and their child’s electronic medical record to be accessed. Parents provided written consent for their child to be included in the study. Identifiers presented in plots and data are deidentified and do not represent patient or research subject identifiers.

Demographic variables collected included age (months), race, and sex. Children who utilized intermittent catheterization to empty their bladder or who had neurogenic bladder were excluded. Clinical variables included the number of confirmed UTIs using the American Academy of Pediatrics definition for UTI, vesicoureteral reflux status (VUR) and history of antibiotic use.[23] Chart review from our institution was used to identify historical UTIs, along with pediatric office records and outside facility records, if an interview with the family indicated a UTI may have occurred outside our institution. These records were then examined to verify if they met the definition of UTI from the American Academy of Pediatrics.

### Specimen collection

Urine specimens were collected from catheterized urine (minimum volume 10 mL) into a sterile urine specimen cup before introducing iodinated contrast for the VCUG procedure. Urine was processed within 24 hours of collection via vacuum filtration on 0.22 µm filters. The urine biomass, collected on these filters, was stored in a –80°C freezer until all samples were ready for DNA extraction.

### DNA extraction, preparation, and sequencing

DNA was isolated from 0.5” x 0.5” fragments of the filters. 1ml of phosphate-buffered saline (PBS) was used to wash the biomass off the filter. DNA isolation was performed using an optimized version of the QIAamp Fast DNA Stool Mini Kit (Cat No./ID: 51604) protocol supplemented with 60 mg/mL lysozyme (Thermo Fisher Scientific, Grand Island, NY).[24–26] The DNA samples were quantified (Qubit Quant-iT PicoGreen kit) and divided into aliquots and then sequenced in triplicates by LoopGenomics (now – Element Biosciences). The LoopGenomics technology uses unique molecular identifiers to build a scaffold of high-quality Illumina short reads that cover the full length of the 16S rRNA gene.[22] This approach minimizes biases found in traditional amplicon sequencing, such as amplification bias of certain taxa and amplification of contaminants. This is especially significant in samples of low biomass, which is expected in children’s urine samples. PBS-treated filters were used as negative controls, and a mock community dilutions series as positive controls to determine background noise and detection limits.

### Microbiome data processing

The sequencing data was processed and summarized into bacterial taxa. Raw sequences were processed using DADA2, following recent recommendations for modifications to accommodate synthetic long reads.[22, 27] Taxonomy was assigned using Bayesian LCA-based Taxonomic Classification Method (BLCA) with the NCBI 16S microbial database (downloaded on 11/5/2021).[28] To broadly assess the characteristics of microbial communities, we used a suite of analyses designed for 16S rRNA sequencing available in R, primarily in the phyloseq and vegan packages.[29] The evenness and richness of each individual’s urinary microbiome were summarized using alpha diversity metrics (number of observed species/genera, the Shannon Index, and Inverse Simpson Index). The alpha and beta diversity measures were compared between groups using standard statistical tests (non-parametric Wilcoxon rank sum for alpha diversity and PERMANOVA for beta diversity). Differences in relative abundance of individual taxa were assessed between groups when the distribution across the dataset was sufficient.

## Results

Fifty-four urine samples were collected, with 31 (57%) from females and 23 (43%) from males (**Table 1**). The mean age of the children was 40.7 months (range 3-130 months). Age was significantly different between sexes, with females being, on average 51.4 months and males being 26.2 months (p = 0.001). Due to this, for the between sex analysis, we limited the data to individuals less than 60 months of age. Thirty-two percent of children in the study reported greater than 3 UTIs, 48% had 1-2 UTIs, and 17% had never had a UTI. Sixty-five percent were currently using a prophylactic antibiotic to reduce their risk of UTI at the time of urine collection, 9% had never used an antibiotic for any reason, and the remaining had intermittently used antibiotics in the past as treatment or prophylaxis. The VUR status at the time of sample collection was 65% (36) had confirmed VUR, 11% (6) had a history of VUR that had resolved, and 24% (13) had never had VUR.

**Table 1.**
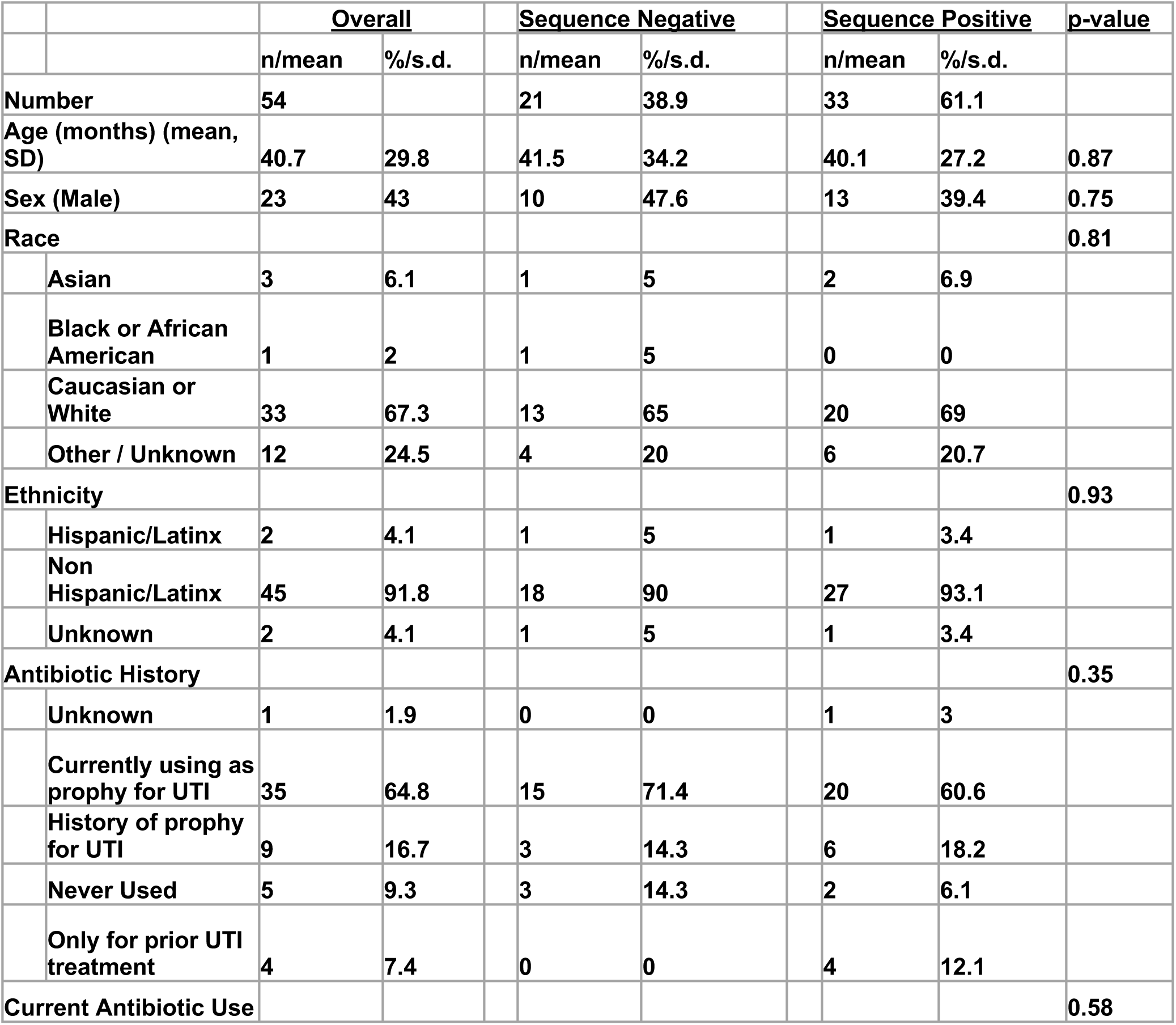
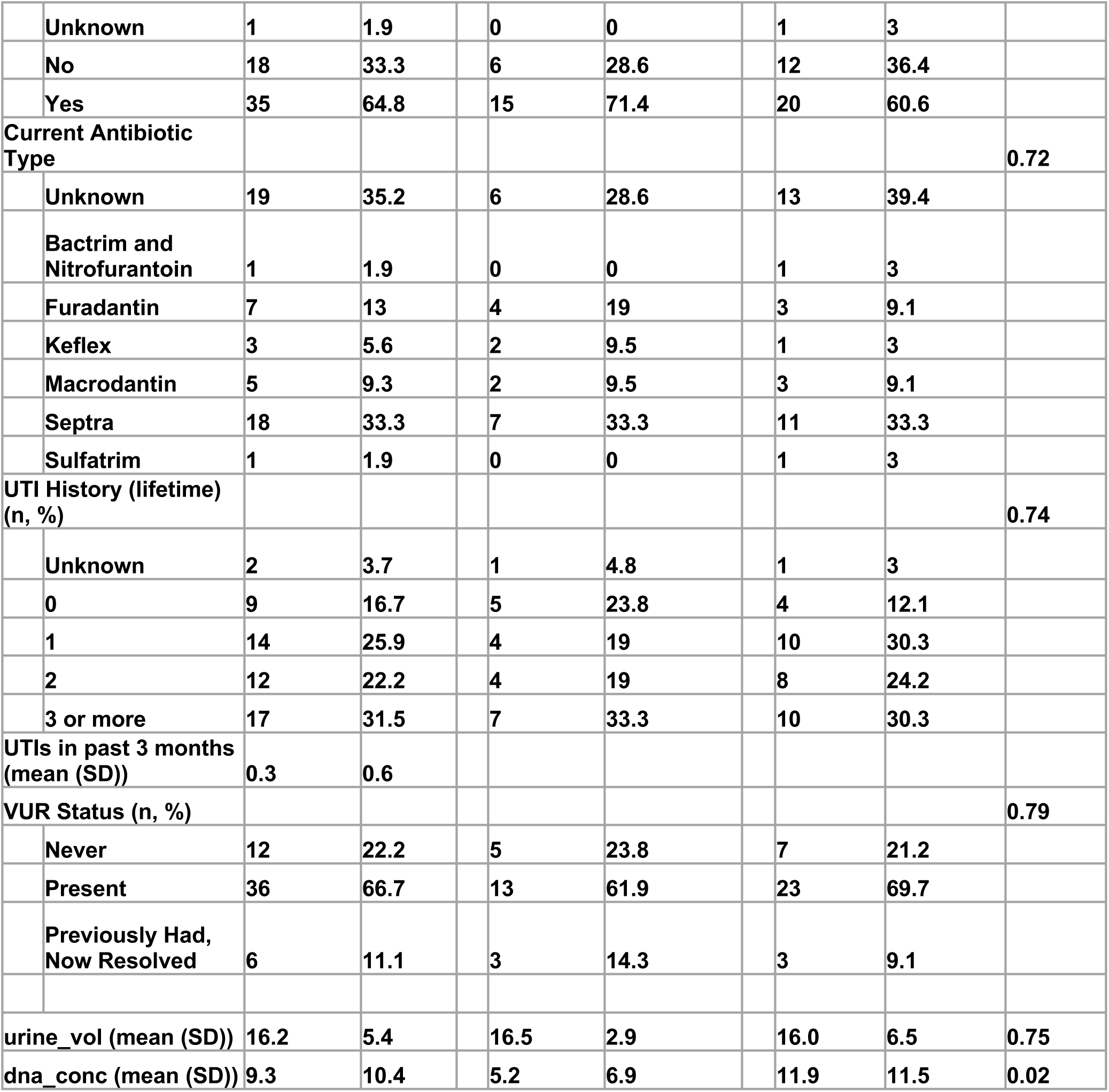
Participant Demographics. The overall cohort is described, as well as sequence negative and sequence positive subsets. All data are reported as number of participants (n) and precents (%), unless noted as mean (standard deviation). There were no significant differences found between participants who had detectable bacteria in their urine (sequence positive) and those who did not (sequence negative) in terms of general demographics, antibiotic use history or UTI history. There was a significant difference in the amount of DNA present in the urine specimen.

### Feasibility of synthetic long-read sequencing of bacteria in pediatric catheterized urines

Sequencing resulted in 0 – 318,342 reads per sample, with a mean of 17,207 reads. All negative controls failed to produce sequences. To determine the minimum acceptable sampling depth per sample, the data were randomly subsampled to various sampling depths ranging from 100 to 10,000. The subsampled data were evaluated based on the relative abundance of individual taxa and alpha diversity measures. We noted no significant differences between these measures of the unrarefied and rarefied data at various thresholds down to 500 reads per sample (**Fig. S1**). We additionally evaluated our mock microbial positive-control samples, which showed the expected composition at the 500 reads per sample threshold. Therefore, we used the threshold of 500 reads per sample as the minimum number of reads to declare a sample as sequence positive and as a subsampling depth for analysis. 33 of the 54 urine samples (61%) provided sequencing data at this threshold and were deemed sequence-positive and subject to further analysis. The overall summary for sequence-positive and sequence-negative samples is given in **Table 1**. There were no significant associations between demographic or clinical variables in individuals whose samples were sequence-positive versus sequence-negative. The sample DNA concentration was significantly increased in the sequence positive samples (5.23 +/- 6.28 ng/ml in sequence negative; 11.85 +/- 11.47 ng/ml in sequence positive samples, *p* = 0.02).

Each DNA sample was aliquoted for sequencing in triplicate and demonstrated little variability compared to that seen across samples. Stacked bar plots representing the relative abundance of bacteria in triplicate of four samples are shown in **Figure 1A**. Overall variability across different taxa in each triplicate set is also low (**Fig. 1B**).

**Figure 1.**
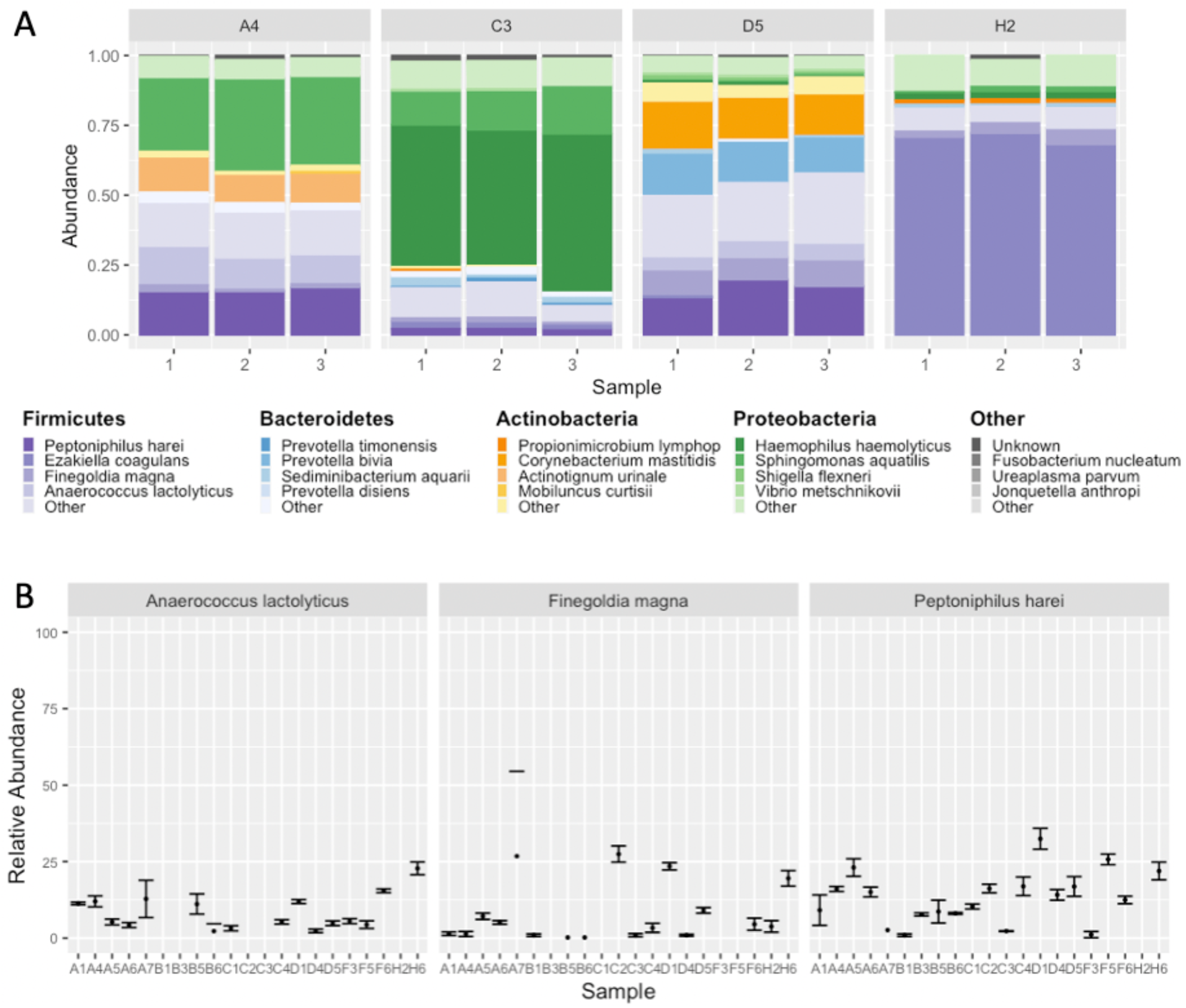
Reproducibility of the synthetic long reads for pediatric urine samples. Each sample was sequenced in triplicate. **(A)** Stacked barplots demonstrating overall similarity between replicate samples. Each sample (facet) was processed in triplicate (x-axis) and demonstrated little variability compared to that seen across samples. **(B)** Variability of relative abundance of select bacterial species across replicates. Values shown are average +/- standard deviation.

All additional analyses were performed using a single replicate of each of the 33 sequence-positive samples.

### Residential bacterial DNA detected in majority of pediatric catheterized urines

Samples from the female and male sexes had polymicrobial urinary microbiomes composed of several bacteria (**Fig. 2A**). We detected an insignificant increase of unique species in females (median 31 species, IQR 18) compared to males (median 16.5 species, IQR 14.25, p-value 0.11). Overall, there was high variability amongst individuals (3 to 42 species, and 2 to 36 genera per sample).

**Figure 2.**
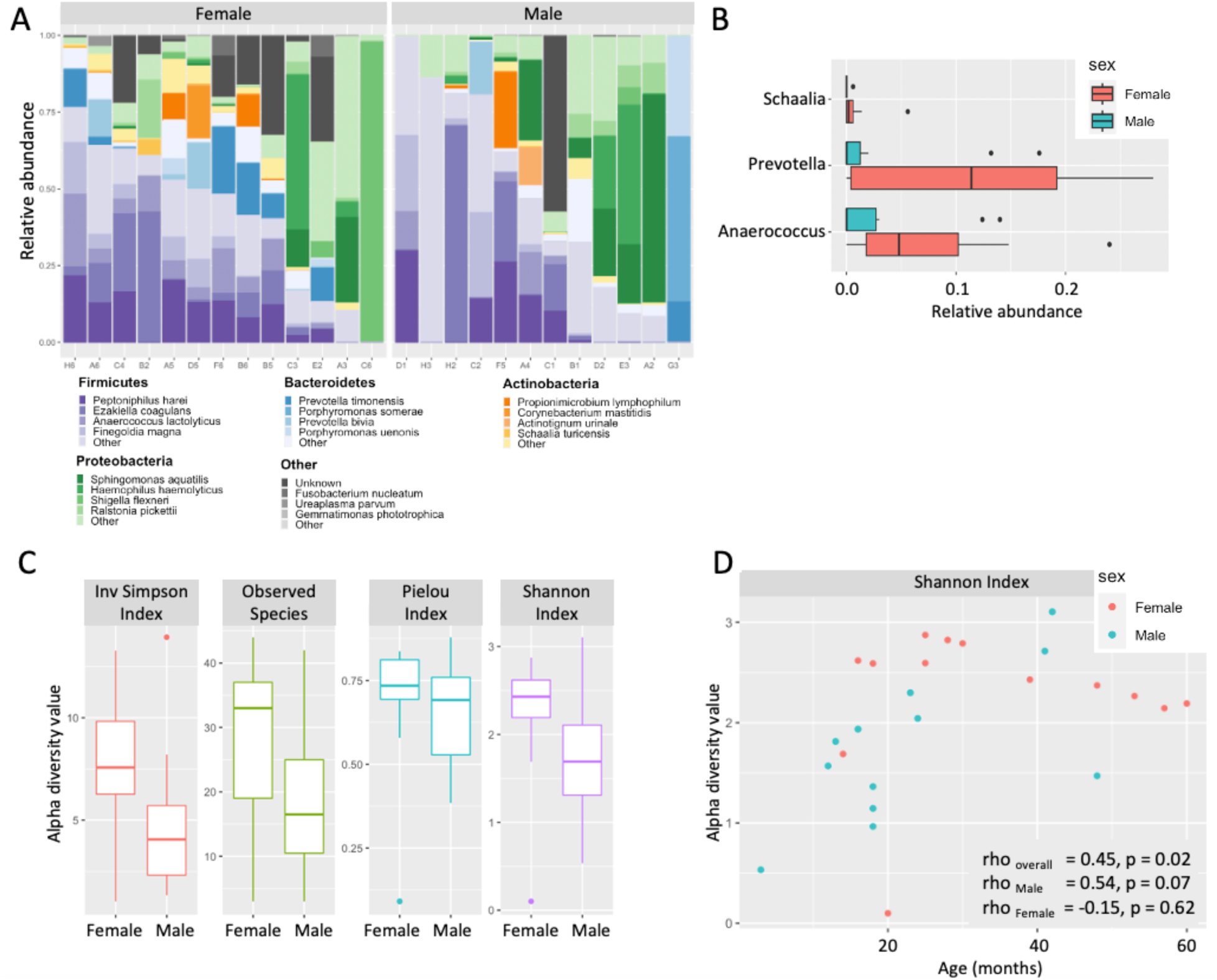
The pediatric urobiome composition varies by sex and age. We observed that the pediatric urobiome is diverse between individuals (A), but there are some overall differences in specific taxa between males and females (B). Alpha diversity also demonstrates differences by sex, with significant differences in the Observed and Shannon Indices (p -, C). Diversity also has a significant positive correlation with age, which is mostly attributed to males (D).

In boys, the dominant taxa included *Peptoniphilus, Ezakiella, Sphingomonas, Ralstonia,* and *Anaerococcus* (**Fig. S2**). In girls, the most abundant were *Prevotella, Peptoniphilus, Anaerococcus, Ezakiella*, and *Streptococcus* (**Fig. S2**). These predominant taxa do not include taxa commonly associated with adult male and female urobiomes – *Lactobacillus* in females and *Staphylococcus* in males. These taxa were present in a small number of samples (2 for *Lactobacillus*, 8 for *Staphylococcus*), in low abundances (maximum 1.0% for *Lactobacillus*, 6.8% for *Staphylococcus*), and not associated with a specific sex. Of note, *Anaerococcus* and *Prevotella* were significantly increased in females compared to males (p-value 0.03 and 0.04, respectively; **Fig. 2B**). Additionally, *Schaalia* was also significantly increased in females (p = 0.02), though it was only detected in 10 samples (9 female samples, one male sample) at low relative abundance (maximum 5.6%). These genera were primarily attributed to the specific species of *Prevotella timonensis*, *Schaalia turincensis*, and *Anaerococcus lactolyticus*. We detected a low abundance of *Actinotignum schaali* in 10 samples, in which 9 were females.

### Urobiome diversity is higher in girls and increases with age in boys

Overall diversity was higher in samples from the females as measured with inverse Simpson (p=0.05) and Shannon (p=0.04) metrics but not by the pielou evenness (p = 0.35) or observed species (p = 0.11) (**Fig. 2C**). Increased age was also associated with increased diversity (rho = 0.45, p = 0.02, **Fig. 2D**). In samples from females the diversity is largely unchanged across age (rho = −0.15, p = 0.62), whereas, in males, diversity increases with age (**Fig. 2D**, **Table 1**) with a correlation factor of 0.54 and p = 0.07.

### Recurrency of UTI results in reduced urobiome diversity

There was variation in the number of UTIs a child had experienced to date (**Table 1**). When grouped by their history of UTI, there were significant differences in the Inverse Simpson index (p = 0.03), the Shannon index (p = 0.05), and Pielou index (p = 0.01). The group with a history of 3 or more UTIs demonstrated significantly reduced urobiome diversity compared to those with a history of only 1 UTI (**Fig. 3A**). The four children who had never had a UTI were excluded from this analysis. When individual taxa were compared among the groups of patients with 1, 2, or 3 or more UTIs, several taxa were decreased in individuals with 3 or more UTIs compared to those with 2 or fewer (**Fig. 3B**). Those are genera of *Enterococcus (significant decrease between 2 UTIs and 1 UTIs, p = 0.05), Lawsonella (significant decrease between 3+ UTIs compared to 1 UTI, p = 0.05; decrease between 3+ UTIs compared to 2, p = 0.07), and Corynebacterium (significant decrease between 1 UTI compared to 3 +, p = 0.05; decrease between 2 UTIs compared to 3+, p = 0.07)*. At the Phyla level, we noted a decrease in Bacteroidetes and an increase in Proteobacteria in children with had 3+ UTIs, however, these changes did not reach significance (p=0.17, p=0.11, respectively), likely due to our small sample size.

**Figure 3.**
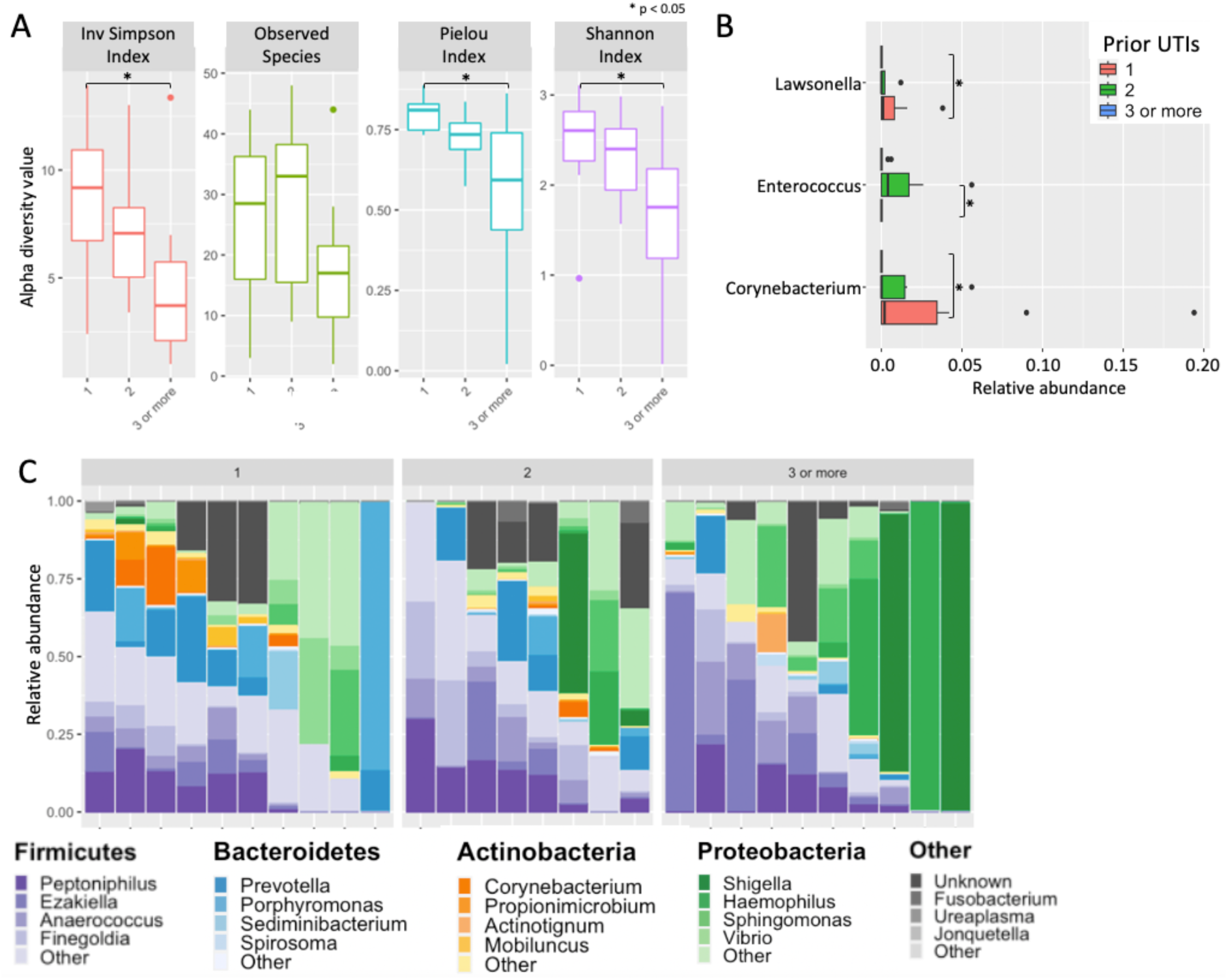
The pediatric urobiome composition varies by UTI history. (A) We observed that the pediatric urobiome is overall less diverse in children with a history of 3 or more UTIs in terms of diversity, with significant decreases in diversity as measured by the Inverse Simpson, Shannon, and Pielou indices between children with 3 or more UTIs compared to those with history of only 1 UTI (p = 0.03, p = 0.05, and p = 0.01, respectively). (B) Three genera demonstrated differences based on UTI history: Lawsonella (significant decrease between 3+ UTIs compared to 1 UTI, p = 0.05; decrease between 3+ UTIs compared to 2, p = 0.07); Enterococcus (significant decrease between 2 UTIs and 1 UTIs, p = 0.05); and Corynebacterium (significant decrease between 1 UTI compared to 3 +, p = 0.05; decrease between 2 UTIs compared to 3+, p = 0.07). (C) Relative abundance of bacterial phyla amongst cohorts of children with history of 1, 2, 3+ UTI.

## Discussion

Results presented here show that even in the low biomass catheter-collected urinary samples of children, we can obtain species-level compositional data without using full shotgun metagenomic sequencing. Additionally, this method expands previously successfully applied species-resolution methods (EQUC and standard urine culture) to obtaining species-level information for unculturable urinary constituents.

Prior studies established that adult urobiomes differ significantly by sex and have different predominant taxa.[30–33] It is not clear whether these sex differences develop early in infancy due to variable anatomy and early hormonal surges or later in childhood due to the hormonal changes of puberty or a combination of these factors. From the microbial ecology perspective, the bladder represents a unique set of conditions and selective pressures that make it clearly distinct from the surrounding environments of skin niches, vaginal and gut microbiomes. These environmental pressures (low oxygen, fluctuating pH and nutrient composition, high concentration of chemicals like urea) may define urobiome composition. In the absence of hormonal changes, one might hypothesize that female and male sex urobiomes may be compositionally close before puberty. The data we obtained contradicts such a hypothesis, showing a distinction between female and male sex urobiome prior to puberty hormonal changes. Interestingly, there are several taxa that appear to be common at high abundances between the two sexes: *Peptonephilus* and *Anaerococcus*.

There are unique sex hormonal differences that occur before birth and within the first year of life, but in our cohort, we did not have representation of infants below one year of age. In the next four years of life, we observed increased urobiome diversity that was driven by the male sex’s samples. This observation correlates with the hypothesis that the different lengths of the female and male urethra may influence the rate of diversity change after birth. The shorter female sex urethra may allow for early population of the urobiome, whereas the longer male sex urethra slows the rate of developing a diverse urobiome. This may imply that the bladder niche has some equilibrium or overall capacity for diversity.

Prior studies from Storm and colleagues did not observe age-related change, which is likely due to collapsing the overall age group of 0-3 year old children and comparing those with the older cohort only.[16] We did not observe the changes in the female sex’s urobiome diversity observed during puberty that was found in that study. This was likely due to the lack of a comparable older age group of females. In a sample of 85 children under 48 months of age, Kinneman et al. found that diversity increased with age.[14] This is confirmed in our results. Our results also support Fredsgaard et al.’s results from 30 clean catch voided urine samples of prepubertal children where females had higher richness and diversity in their urinary microbiome than males.[18]

Considering the composition in the pediatric samples we presented, some prominent genera from the adult urobiome are not prominent in children. For example, in adult males Corynebacterium and Staphylococcus are frequently abundant in the urobiome.[34–36] However, we did not detect these genera robustly in male samples; they were only identified in a few samples at low abundance except for in one instance. Moreover, genera previously classified as Lactobacilli[38] that are the most common genera in adult female sex urobiomes were not found robustly in our dataset, with only 2 samples having a Lactobacillus genera, each at a low abundance. This is consistent with the observations made by Storm et al. from their data that Lactobacilli appear in female populations during puberty.[16] There are abundant taxa that appear to be present in early childhood that may also remain prevalent in adult bladders, for example, *Prevotella, Streptococci,* and *Porphyromonas*. Our population in this study was specific and consisted of children referred for a VCUG due to urologic concerns with the majority having a history of VUR. Results from this data may not be fully generalizable to larger populations of healthy children, but the aims of this study were still achieved. Due to this, 60.6% of all sequence-positive samples (**Table 1**) came from children who were taking prophylactic doses of antibiotics as a measure of preventing recurrent febrile UTIs. Surprisingly, prophylactic use of antibiotics did not result in the elimination of the urobiome. Males below 1 year of age are more susceptible to UTI.[39] Around 1 year of age, females become more at risk for UTI compared to males, up until the late stages of life when the immune system of both sexes weakens due to senescence. We observed early male urobiome had low bacterial diversity. However within our cohort we did not have males with ‘sequence-positive’ samples under one year of age.

Comparing patient cohorts who have had different numbers of UTIs showed that the diversity was reduced in those individuals who had 3 and more UTIs. There are several possible explanations for such observations, such as alterations of the urobiome due to antibiotic exposure and changes associated with prior overgrowth of uropathogens during the UTIs. In our dataset, only 2 samples were ‘sequence-positive’ from children who had never used antibiotics, while all other ‘sequence-positive’ samples were from children who were either currently taking prophylactic antibiotics or were treated with antibiotics previously. We do not have enough statistical power to establish significance.

Still, there was no difference observed in the diversity of the urobiome between those with prior antibiotic use and current prophylactic antibiotic use. This observation suggests that if there is a reduction of the diversity or other parameters in the urobiome due to antibiotic treatments (including current prophylactic use), the changes might be smaller than those associated with UTI-caused shifts.

Our pilot study has some limitations, including the use of convenience sampling and a small number of subjects. We did not capture the circumcision status of male sex children; recent studies have identified a microbiome on the foreskin, which may have been detected in catheterized urine, especially in younger children with physiological phimosis whose foreskin can’t be adequately retracted for catheterization directly into the urethra.[30] We did not run short amplicon sequencing in parallel with LoopSeq due to concerns of the limited amount of DNA extracted. This study also had strengths, including using a filtering technique that does not utilize a high-speed centrifuge, potentially expanding the number of centers where urine processing could be done. We additionally used synthetic long-read sequencing that enabled species-level identification. Such methodology balances access and cost between newly established methods for obtaining compositional data for urine samples for microbiome analysis.

In summary, our study confirms prior observations that even infants have complex urobiomes. In our cohort, we identified the urobiome in children as young as 3 months and obtained species-level taxonomic resolution by using culture-independent full-length 16S sequencing. We showed that the urobiome composition varies amongst individuals and demonstrates notable changes between sexes and with age. We identified specific species overrepresented in females and patients with repeated UTIs. With observed changes in urobiome diversity levels, our study suggests a connection between decreased overall urobiome diversity and recurrent UTIs in children. Overall, our pilot results show the feasibility of the species-level urobiome investigation in future broader cross-sectional as well as longitudinal studies exploring urobiome stability within an individual, changes with health status alterations, and drug applications.

## Data Availability

Data produced in the present study are available upon reasonable request to the authors

