## Supplemental Information for "Characterization of pediatric urinary microbiome at species-level resolution indicates variation due to sex, age, and urologic history"

**Supplemental Table S1.** Detailed summary of pediatric urobiome studies to date. The collection methods used were either transurethral catheterization (TUC) or clean-catch midstream urine (CCMSU). Patients ranged from those with a healthy urinary tract (UT), those suspected of having urinary tract infections (UTI) in the emergency room (ER), and those with UT pathologies such as neuropathic bladder or vesicoureteral reflux (VUR).

| Study | Sample Sex and Age | Characteristics | Collection | EQUC | 16S Region |
|---|---|---|---|---|---|
| Forester et al, 2020 | Mixed, 2 months – 252 months | Neuropathic bladder | TUC, > 5mL | | V4 |
| Fredsgaard et al, 2021 | Half male/female, prepubertal on Tanner scale | Asymptomatic, healthy UT and no use of antibiotics within 3 months | CCMSU, 1 sample divided into 2 aliquots of 10 mL | | V4 |
| Hadjifrangiskou et al 2023 | Males, infants (< 12 months) | Elective circumcision, healthy UT, and no prior UTI | TUC | Yes | V4 |
| Kassiri et al 2019 | Males, 3 months – 96 months | Antibiotic/ prophylaxis usage vs. no usage | TUC prior to elective urologic procedures, 15 mL | | V6 |
| Kinneman et al 2020 | Mixed, < 48 month | Suspected UTI in ER visit but no other UT pathologies | TUC due to diagnosing UTI, 1 mL | | V3/V4 |
| Storm et al 2022 | Mixed, 0-18 years | Under anesthesia for procedures, no use of antibiotics within 3 months, and healthy UT | TUC | Yes | V4 |
| Vitko et al 2022 | Mixed, pediatric + 9 adult | Healthy controls, VUR patients (with and without renal scarring) | TUC or CCMSU for controls, TUC for VUR | | V4 |

**Supplemental Figure S1. Evaluation of sampling depth.** The data from each sample were randomly subsampled to 100, 500, 1,000 and 10,000. The subsampled data were evaluated on the basis of the relative abundance of individual taxa and alpha diversity measures. A) Alpha diversity for four representative samples are displayed in A, demonstrating little variability at sampling depths of 500 reads or greater. B) stacked bar plots of representative samples demonstrating little variability across subsampling depths. **Note:** Samples 38 and 45 did not have enough read depth to sample at 10,000 reads.

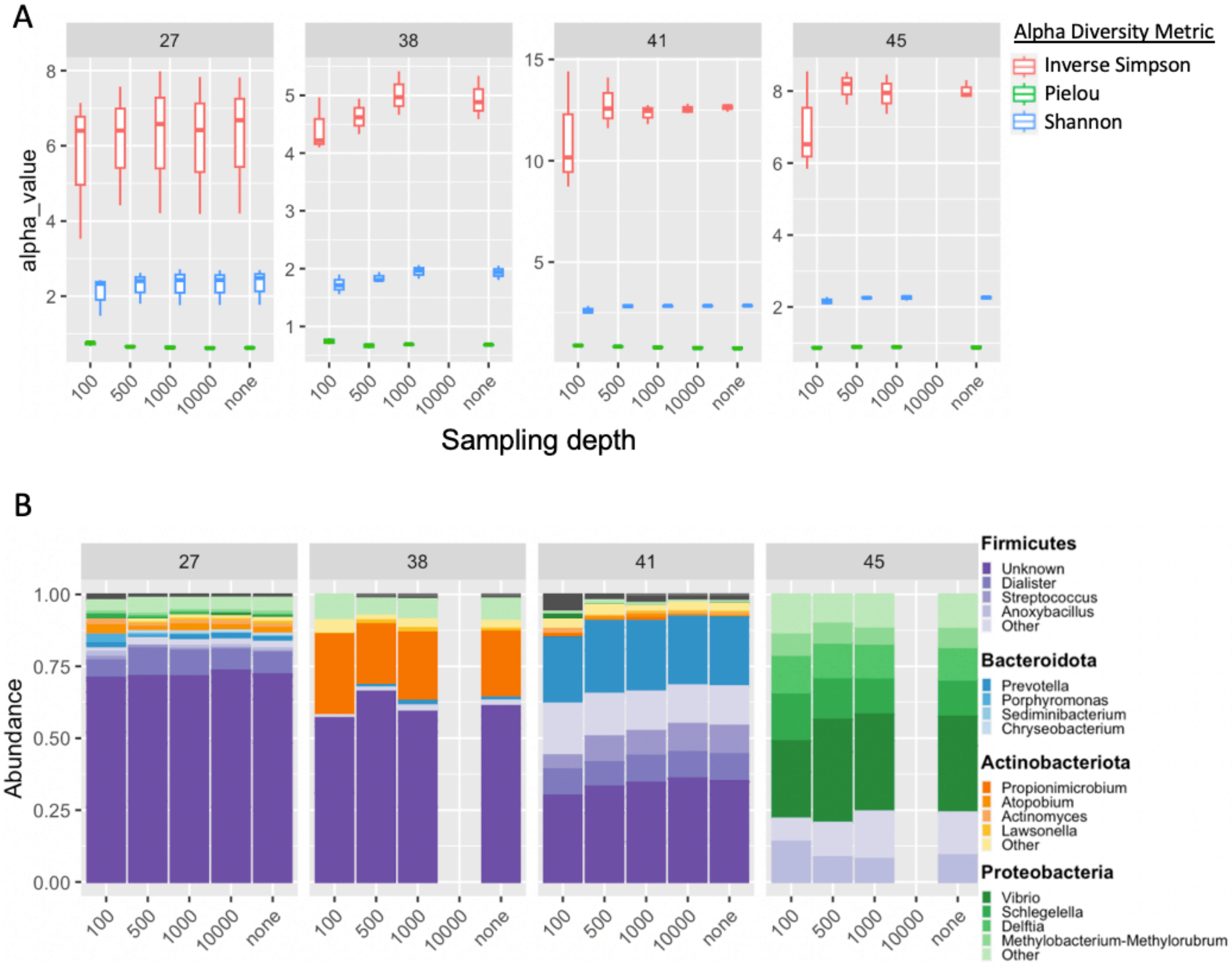

**Supplemental Figure S2.** Top genera in pediatric urinary microbiome varies by sex. Boxplots representing the median and interquartile range of individual bacterial genera for females (left) and males (right).

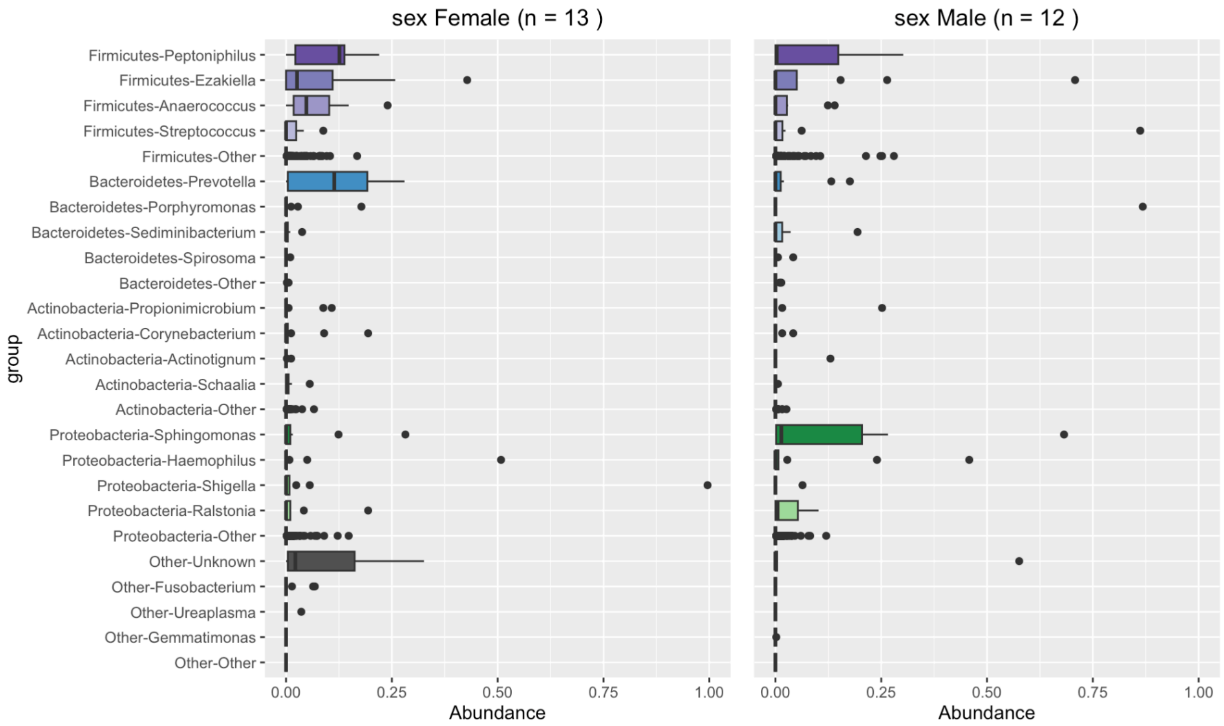
